# Patterns of social-affective responses to trauma exposure and their relation to psychopathology

**DOI:** 10.1101/2023.07.25.23293160

**Authors:** Sarah Thomas, Judith Schäfer, Philipp Kanske, Sebastian Trautmann

**Affiliations:** Institute of Clinical Psychology and Psychotherapy, Faculty of Psychology, Technische Universität Dresden; Department of Psychology, Faculty of Human Science, Medical School Hamburg; ICPP Institute of Clinical Psychology and Psychotherapy, Medical School Hamburg; Max Planck Institute for Human Cognitive and Brain Sciences, Leipzig, Germany

## Abstract

**Introduction:** Traumatic event exposure is an important risk factor for the development and maintenance of psychopathology. Social-affective responses to trauma exposure (e.g. shame, guilt, revenge, social alienation) could moderate this relationship, but little is known about their relevance for different types of psychopathology. Moreover, the interplay of different social-affective responses in predicting psychopathology is poorly understood.

**Methods:** In a sample of *N*=1321 trauma-exposed German soldiers, we examined cross-sectional associations of trauma-related social alienation, revenge, guilt and shame with both categorical (depressive disorder, alcohol use disorder, posttraumatic stress disorder) and dimensional (depression, anxiety) measures of psychopathology. Latent class analysis was conducted to identify possible patterns of trauma-related social-affective responses, and their relation to psychopathology.

**Results:** All trauma-related social-affective responses predicted the presence of posttraumatic stress disorder, depressive disorder, alcohol use disorder and higher depressive and anxiety symptoms. Three latent classes were identified that fitted the data best, reflecting groups with (1) low, (2) moderate and (3) high risk for social-affective responses. The low-risk group demonstrated the lowest expressions on all psychopathology measures. Compared to the moderate-risk group, the high-risk group demonstrated no increased psychopathology.

**Conclusions:** Trauma-related social alienation, shame, guilt, and revenge are characteristic of individuals with posttraumatic stress disorder, depressive disorder, alcohol use disorder, as well as with higher anxiety and depressive symptoms. There was little evidence for distinctive patterns of social-affective responses despite variation in the overall proneness to show trauma-related social-affective responses. Trauma-related social-affective responses could represent promising treatment targets which might be included in both cognitive and emotion-focused interventions.

## Introduction

Exposure to traumatic events is an important risk factor for the development and maintenance of mental disorders (1). Apart from Posttraumatic Stress Disorder (PTSD), trauma exposure is particularly associated with the development of major depressive disorder and alcohol use disorder (AUD) (2). However, individuals vary considerably in their response to trauma exposure and the majority of individuals adjust well to the experience of severe stressful or traumatic events (3). Numerous factors have been suggested to moderate the association between trauma exposure and psychopathology (4). Social factors, which have received less attention so far, are among those variables that could have a decisive influence on mental health after trauma exposure (5). Relevant social factors include for instance perceived social support, disclosure and social acknowledgement (5).

Among social factors, social-affective responses to trauma exposure could be of particular importance. Following the socio-interpersonal model of PTSD by Maercker and Horn (6), social-affective responses to trauma exposure can be understood as complex mental states encompassing feelings, cognitions and motivations that relate to the social reality of an individual. Social-affective responses to traumatic events can include positive responses such as compassion (7) but can also include negative responses, such as shame, guilt, revenge and social alienation (6, 8). In line with the socio-interpersonal model of PTSD, most authors conceptualize guilt (9), revenge (10), shame and social alienation (8) as complex states that are relevant from both a cognitive and an emotion-based perspective of posttraumatic processing. Cognitive models of posttraumatic stress assume that dysfunctional trauma appraisals lead to negative cognitive schemas about the self and the world and produce a sense of ongoing threat accompanied by diminished self-efficacy (11, 12). In this context, trauma-related shame, guilt, and social alienation, for example, have been considered both as elements and consequences of negative cognitive schemas about the self and the world (11, 12). From an emotion-based perspective, shame and guilt, and in some interpretations also feelings of estrangement and vengefulness (6), are conceptualized as social emotions (13). Social emotions are regarded as “cognition-dependent” emotions that require mental representations of both oneself and others and work in the service of a social goal (14). Recent theories and empirical findings increasingly emphasize the importance of distressing social emotions as possible responses to trauma exposure (13). Importantly, negative trauma related social-affective responses could be important for posttraumatic processing beyond general trauma-related emotional distress and negative cognitions. Social-affective responses such as shame, guilt, or social alienation may be particularly difficult to manage because they can threaten a person’s sense of self and social identity (15) and could seriously affect social relationships by preventing individuals from perceiving and using potential social resources such as social support or group membership (16). Moreover, there is evidence that social affective responses such as shame keep individuals from seeking professional help (17).

In line with these assumptions, negative social-affective responses to trauma exposure have been associated with higher levels of psychopathology in previous studies (5). Trauma related guilt and shame have been investigated most frequently and are associated with higher levels of PTSD symptoms (18, 19), with some authors suggesting a model of guilt and shame-based PTSD (15). Posttraumatic guilt and shame are highly interrelated, but it is assumed that the relationship between guilt and PTSD is more variable and less strong than the relationship between shame and PTSD (18, 19). Besides shame and guilt, trauma-related social alienation has shown to be an important mediator of the association between trauma exposure and PTSD symptoms (20). Trauma-related revenge phenomena have received less attention so far, although posttraumatic revenge feelings and cognitions have found to be predictive of higher severity and maintenance of PTSD symptoms (10, 21). To date, social affective responses to trauma exposure have mainly been investigated with respect to PTSD. However, there is also some evidence regarding other forms of posttraumatic psychopathology, such as depressive symptoms. Similar to PTSD, trauma-related shame (22) and guilt (22, 23) have been associated with higher levels of depressive symptoms. Like for PTSD, there is evidence that trauma-related social alienation mediates the association between traumatic event exposure and depressive symptoms (20). Moreover, posttraumatic guilt and shame have been related to increased alcohol use and might be associated with higher levels of anxiety symptoms (22, 24).

Taken together, negative social-affective responses to trauma exposure have been associated with higher levels of subsequent psychopathology. Previous studies have focused primarily on PTSD and less is known about associations with other psychopathologies such as depressive disorder (DD) and AUD. In addition, most studies have examined shame and guilt, while other possible social-affective responses have received less attention. Moreover, the interplay of different social-affective responses in predicting mental health has rarely been studied. Thus, little is known about whether there could be distinct patterns of different social-affective responses to trauma exposure and whether they relate differentially to psychopathology.

We therefore aimed to investigate associations of negative social-affective responses to trauma exposure (social alienation, revenge, guilt, shame) with mental disorders (DD, AUD and PTSD) as well as with dimensional measures of depression and anxiety in trauma exposed individuals. We also aimed to investigate whether there are distinguishable patterns of trauma-related social-affective responses and, if so, how these patterns relate differentially to mental disorders and to dimensional symptom measures.

## Methods

### Participants and procedure

Data were collected between 27.04.2010 and 10.12.2010 as part of the cross-sectional component of a larger study program (25) investigating mental health and its determinants in German military personnel. A comprehensive description of the entire study design can be found elsewhere (25). A total of *N* = 2372 German soldiers were included in the original study. For the purpose of the present analysis, only participants who had been exposed to at least one lifetime traumatic event according to the DSM-IV A1 criterion were included (*N* = 1636). Since the low proportion of females in the German military would not have permitted adequate subgroup analysis, female soldiers (*n* = 104) were excluded in the present analysis. Moreover, participants who had any missing values on the items measuring shame (*n* = 207), guilt (*n* = 206), revenge (*n* = 206) and social alienation (*n* = 204) were excluded. This resulted in an analysis sample of *N* = 1321 individuals. To ensure that there was no selective non-response in the sense that more distressed individuals did not respond to the items, we examined whether the excluded participants (*N* = 211) and the analysis sample (*N* = 1321) differed with respect to the outcomes examined. There were no differences regarding the severity of depressive and anxiety symptoms and regarding the percentage of PTSD and AUD, but excluded individuals had a lower percentage of DD than included individuals (Table S1).

Participation in the study was voluntary and confidential. Trained clinical psychologists completed informed consent procedures and conducted the assessments. Informed written consent was obtained from all participants. Assessments comprised a standardized diagnostic interview with supplementary questionnaires to allow for the collection of additional information, such as dimensional symptom severity and demographic data. The study was approved by the Ethics Board of Technische Universität Dresden (EK 72022010).

### Measures

#### Social-affective responses to trauma exposure

Social-affective responses to trauma exposure (shame, guilt, revenge, social alienation) were measured with items adapted from the a priori item pool of the Posttraumatic Cognitions Inventory (PTCI) that were of interest to the present research question (11, 26). Social-affective responses (past four weeks) were assessed with respect to the worst traumatic event. Items were rated on a 5-point scale (“Strongly disagree”, “rather disagree”, “neutral”, “rather agree”, “strongly agree”). Since several response categories had too low counts to treat the variables as dimensional, they were operationalized as dichotomous variables (present vs. not present). As shown in the online supplement (Table S2) only a very small percentage of participants agreed to the items. Given the male military sample, it is possible that emotional and potentially stigmatizing constructs such as shame, guilt, revenge, and social alienation were underreported (27). Therefore, the middle response (“neutral”), which can be conceptualized as transition point between disagreement and agreement in Likert-type scales, was chosen as a cut-off for the presence of the respective social-affective response.

Guilt was defined as feelings and thoughts about having violated personal norms of right and wrong and being responsible for this wrongdoing (i.e. perceived lack of a justification for one’s actions) (15). Guilt was assessed with the item “The way I thought/felt and behaved during the event is unforgivable”. Shame (external) relates to the experience of a negative social presentation and is characterized by feelings and thoughts of being devalued in the eyes of others and being looked down upon (15). We decided to focus on external shame, since external shame has shown tighter links to psychopathology than internal shame (28) and could be easier to distinguish from guilt, as both guilt and internal shame refer to a negative self-evaluation, whereas external shame refers to the perception of being negatively evaluated by others (15). External shame was assessed with two items to be able to consider shame as a response to the actual presence of others during the traumatic event (“I embarrassed myself during the event”) and as a response to the theoretical presence and judgment of others (“If people knew what happened, they would look down on me”). External shame was rated as present if participants responded with at least neutral to either of the two items. We defined revenge as the motivation to retaliate that results from feelings and thoughts of having been hurt wrongfully (29). Revenge was rated as present if the item “I want to punish the people who did this to me” was not negated. Social alienation was defined as feelings and thoughts of being disconnected from others (30). Social alienation was also measured with two items to consider both alienation in close relationships (“I will never be able to be close to other people again”) as well as more generalized appraisals of disconnectedness (“Other people do not understand me”). Social alienation was classified as present if participants responded with at least neutral to either of these items. For shame (0.94) and social alienation (0.95), tetrachoric correlations between the items were high enough to allow the combination of the items into one construct.

#### 12-month mental disorders

The prevalence of a DSM-IV diagnosis of DD, PTSD or AUD in the past 12 months was assessed using the military version of the Munich-Composite International Diagnostic Interview (DIA-X/M-CIDI (31)). The DIA-X/M-CIDI is a fully standardized interview that allows a reliable (32) and valid (33) assessment of mental disorders for lifetime and in the past 12 months according to DSM-IV-TR diagnostic criteria. DD was defined as the presence of either major DD or dysthymia in the past 12 months. AUD included those individuals who had met the criteria of either alcohol abuse or alcohol dependence in the past 12 months.

#### Anxiety and depressive symptoms

Since it was deemed important to consider dimensional measures of psychopathology in addition to the categorical assessment of mental disorders (34), current anxiety and depressive symptoms (past seven days) were assessed with the German version of the Hospital Anxiety and DD Scale (HADS-D) (35). The anxiety and the depression scale of the HADS-D each consist of seven items that are rated on a four point scale. The response scales are anchored differently for each item and measure either the frequency or severity of symptoms or the severity of behavioral changes. A total sum score was calculated for anxiety symptoms (theoretical range 0-21) and for depressive symptoms (theoretical range 0-21). In the present sample, internal consistency was α = 0.75 for the anxiety scale and α = 0.77 for the depression scale.

### Data analysis

All analyses were performed with Stata 15.1 (36). First, logistic regressions were calculated to examine whether and how strongly each individual social-affective response (shame, guilt, revenge and social alienation) predicted the presence of DD, PTSD and AUD, respectively. In order to better assess the specificity of the individual associations, for each logistic regression, an additional model was calculated, adjusting for the respective comorbid disorders of DD, PTSD or AUD. Second, to complement the analyses by dimensional symptom measures, linear regressions were performed to examine individual associations of shame, guilt, revenge and social alienation with depressive and anxiety symptoms. Again, models were re-calculated adjusting for anxiety symptoms in models with depressive symptoms as dependent variable, and vice versa.

Subsequently, Latent Class Analysis was performed to identify potential latent classes of patterns of social-affective responses. The number of classes was determined using the Bayesian Information Criteria and Akaike Information Criteria. In a second step, subjects were assigned to a given class based on their posterior class membership probabilities. To examine whether patterns of social-affective responses were predictive of categorical and/or dimensional measures of psychopathology, logistic and linear regressions were calculated with mental disorders and dimensional symptom measures as dependent and assigned class membership as predictor variable. Models were re-calculated adjusting for anxiety symptoms in models with depressive symptoms as dependent variable, and vice versa. Associations with diagnosis of PTSD, AUD or DD as dependent variable were adjusted for the respective comorbid disorders (PTSD, AUD, DD).

## Results

### Sample characteristics

Participants were male and had a mean age of 28.8 years (*SD* = 7.6). The mean number of experienced traumatic events was 2.6 (*SD* = 1.9). There were 32.0% of participants who had children and 27.8% were married. Among the participants, 18.8% had a low educational level (9^th^ grade), 63.2% had a middle (10^th^ grade) educational level and 18.0% had a high (high school or higher) educational level. Of the participants, 1.7% rated their economic situation as “bad” or “very bad”, 19.8% rated their economic situation to be at least sufficient and 78.5% rated their economic situation as “good” or “very good”. Among the participants. Tetrachoric correlations between shame, guilt, social alienation and revenge are presented in Table 1. High correlations were found between all social-affective responses with the strongest correlation being between guilt and shame (Rho = 0.88). The frequency of the presence of revenge, social alienation, shame and guilt in the total sample and among individuals meeting criteria for PTSD, DD or AUD is shown in Table 2.

**Table 1.**
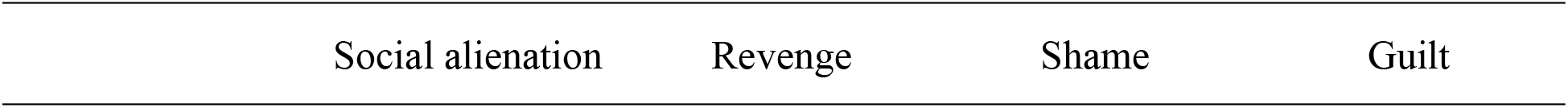
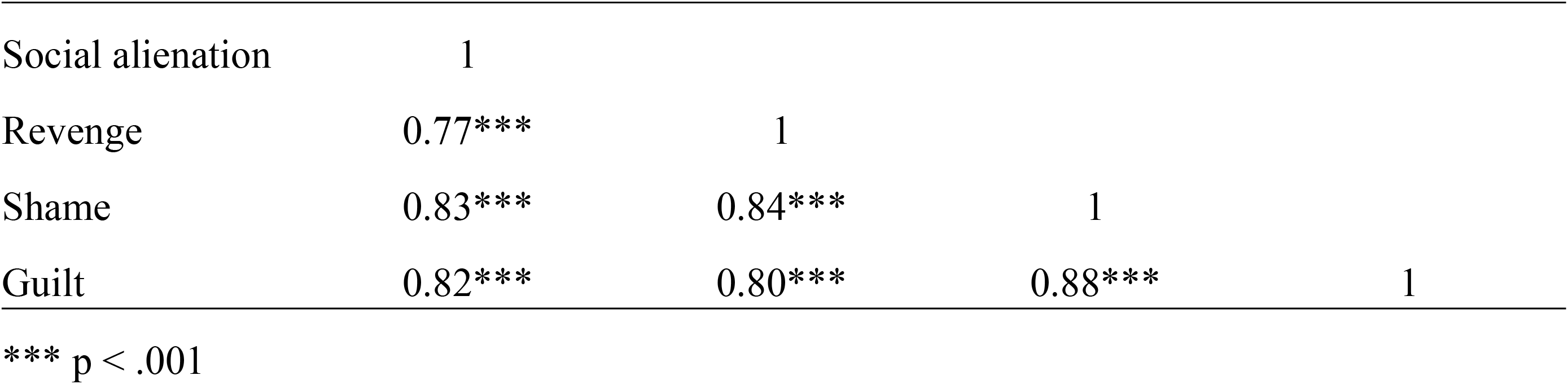
Tetrachoric correlations between social-affective responses to trauma exposure.

|  |  |  |  |  |
| --- | --- | --- | --- | --- |
| Social alienation | 1 |  |  |  |
| Revenge | 0.77*** | 1 |  |  |
| Shame | 0.83*** | 0.84*** | 1 |  |
| Guilt | 0.82*** | 0.80*** | 0.88*** | 1 |

\*\*\* $p < .001$

**Table 2.** Frequency of social alienation, revenge, shame and guilt in individuals with a 12-month diagnosis of PTSD, DD and AUD.

| | Total sample<br>$N=1321$ | DD<br>$N=53$ | PTSD<br>$N=54$ | AUD<br>$N=66$ |
| --- | --- | --- | --- | --- |
|  | n (%) | n (%) | n (%) | n (%) |
| Social alienation | 233 (17.6%) | 24 (45.3%) | 29 (53.7%) | 20 (30.3%) |
| Revenge | 231 (17.5%) | 15 (28.3%) | 23 (42.6%) | 25 (37.9%) |
| Shame | 155 (11.7%) | 14 (26.4%) | 18 (33.3%) | 16 (24.2%) |
| Guilt | 149 (11.3%) | 15 (28.3%) | 12 (22.2%) | 14 (21.2%) |

*Note.* DD = Depressive disorder. PTSD = Posttraumatic Stress Disorder. AUD = Alcohol Use Disorder.

### Association of social-affective responses with mental disorders and with dimensional symptom measures (anxiety and depression)

Table 3 shows the associations of shame, guilt, revenge and social alienation with DD, PTSD and AUD. All associations were statistically significant. The strongest associations existed with respect to PTSD and with respect to social alienation. The highest ORs were observed for associations between social alienation and PTSD (*OR* = 6.04, 95% CI = [3.47, 10.53], *p* < .001) and between social alienation and DD (*OR* = 4.19, 95% CI = [2.39, 7.35], *p* < .001). High ORs were also found for the association between shame and PTSD (*OR* = 4.12, 95% CI = [2.28, 7.46], *p* < .001), revenge and PTSD (*OR* = 3.78, 95% CI = [2.16, 6.61], *p* < .001), and between guilt and DD (*OR* = 3.34, 95% CI = [1.79, 6.23], *p* < .001). All associations were reduced when adjusted for comorbid disorders (Table 3) and there were no statistically significant associations any more between revenge and DD and guilt and PTSD.

**Table 3.** Associations of shame, guilt, revenge and social alienation with DD, PTSD and AUD.

|  | DD |  |  | PTSD |  |  | AUD |  |  |
| --- | --- | --- | --- | --- | --- | --- | --- | --- | --- |
|  | *OR* | *p* | 95%CI | *OR* | *p* | 95%CI | *OR* | *p* | 95%CI |
| **Social alienation** |  |  |  |  |  |  |  |  |  |
| Unadjusted model | 4.19 | < .001 | [2.39, 7.35] | 6.04 | < .001 | [3.47, 10.53] | 2.13 | .007 | [1.23, 3.67] |
| Adjusted model | 3.31 | < .001 | [1.83, 5.98] | 5.06 | < .001 | [2.86, 8.96] | 1.80 | .044 | [1.01, 3.21] |
| **Revenge** |  |  |  |  |  |  |  |  |  |
| Unadjusted model | 1.92 | .037 | [1.04, 3.56] | 3.78 | < .001 | [2.16, 6.61] | 3.11 | < .001 | [1.85, 5.22] |
| Adjusted model | 1.45 | .269 | [0.75, 2.78] | 3.33 | < .001 | [1.87, 5.93] | 2.85 | < .001 | [1.68, 4.83] |
| **Shame** |  |  |  |  |  |  |  |  |  |
| Unadjusted model | 2.87 | .001 | [1.52, 5.42] | 4.12 | < .001 | [2.28, 7.46] | 2.57 | .002 | [1.42, 4.63] |
| Adjusted model | 2.20 | .021 | [1.13, 4.30] | 3.46 | < .001 | [1.88, 6.39] | 2.25 | .009 | [1.22, 4.13] |
| **Guilt** |  |  |  |  |  |  |  |  |  |
| Unadjusted model | 3.34 | < .001 | [1.79, 6.23] | 2.36 | .012 | [1.21, 4.59] | 2.23 | .011 | [1.21, 4.14] |
| Adjusted model | 2.90 | .001 | [1.52, 5.52] | 1.82 | .094 | [0.90, 3.66] | 1.99 | .033 | [1.06, 3.75] |

*Note.* DD = Depressive disorder. PTSD = Posttraumatic Stress Disorder. AUD = Alcohol Use Disorder. Adjusted model: adjusted for the respective comorbid disorders of DD, PTSD or AUD.

Table 4 displays the associations between shame, guilt, revenge and social alienation and anxiety and depressive symptoms. All associations were statistically significantly. As for associations with mental disorders, the highest associations were observed with regard to social alienation. Social alienation predicted higher anxiety (β = 2.02, 95% CI = [1.64, 2.40]), *p* < .001) as well as higher depressive symptoms (β = 1.84, 95% CI = [1.46, 2.21], *p* < .001). A strong association was also found between shame and depressive symptoms (β = 1.60, 95% CI = [1.15, 2.05], *p* < .001). All associations were reduced when adjusted for anxiety and depressive symptoms, respectively (Table 4). The association between guilt and depressive symptoms was not statistically significant any more when adjusted for anxiety symptoms. When adjusted for depressive symptoms, there was no longer a significant association between shame and anxiety symptoms.

**Table 4.** Associations of shame, guilt, revenge and social alienation with depressive and anxiety symptoms.

|  | Depressive Symptoms |  |  | Anxiety Symptoms |  |  |
| --- | --- | --- | --- | --- | --- | --- |
| | $\beta$ | $p$ | 95%CI | $\beta$ | $p$ | 95%CI |
| **Social alienation** |  |  |  |  |  |  |
| Unadjusted model | 1.84 | < .001 | [1.46, 2.21] | 2.02 | < .001 | [1.64, 2.40] |
| Adjusted model | 0.59 | < .001 | [0.28, 0.89] | 0.86 | < .001 | [0.55, 1.16] |
| **Revenge** |  |  |  |  |  |  |
| Unadjusted model | 1.09 | < .001 | [0.70, 1.47] | 1.10 | < .001 | [0.71, 1.49] |
| Adjusted model | 0.39 | .010 | [0.09, 0.69] | 0.39 | .012 | [0.09, 0.69] |
| **Shame** |  |  |  |  |  |  |
| Unadjusted model | 1.60 | < .001 | [1.15, 2.05] | 1.33 | < .001 | [0.87, 1.79] |
| Adjusted model | 0.77 | < .001 | [0.42, 1.12] | 0.27 | .135 | [-0.09, 0.64] |
| **Guilt** |  |  |  |  |  |  |
| Unadjusted model | 1.08 | < .001 | [0.62, 1.54] | 1.35 | < .001 | [0.88, 1.82] |
| Adjusted model | 0.22 | .234 | [-0.14, 0.58] | 0.65 | < .001 | [0.29, 1.01] |

*Note.* PTSD = Posttraumatic Stress Disorder. AUD = Alcohol Use Disorder. Adjusted model: adjusted for depressive symptoms respectively anxiety symptoms.

### Latent class analysis

The fit statistics for different class solutions are displayed in Table 5. The model that fitted the data best was the one assuming three underlying classes of patterns of social affective responses. The three classes model did not differ from a saturated model (χ²(1) = 2.036, *p* = 0.154). The frequencies of shame, guilt, social alienation and revenge within each of the three latent classes of social-affective responses are shown in Figure 1. The majority of individuals (79.2%) were assigned to a *low-risk group* for social affective-responses, 180 participants (13.6%) were assigned a *moderate-risk group* for social affective-responses and 95 participants (7.2%) to a *high-risk group* for social-affective responses. The low-risk group was characterized by no or very low frequencies of social-affective responses. Individuals in this group reported no shame and no social alienation, and only 6.7% of individuals reported revenge and 2.2% reported guilt. In the high-risk group, all individuals confirmed the presence of shame, guilt and revenge and 92.6% confirmed the presence of social alienation. In the moderate-risk group the percentage of individuals reporting guilt (17.2%), shame (33.3%) and revenge (36.7%) was rather low, but a majority (80.6%) reported social alienation.

**Figure 1:**
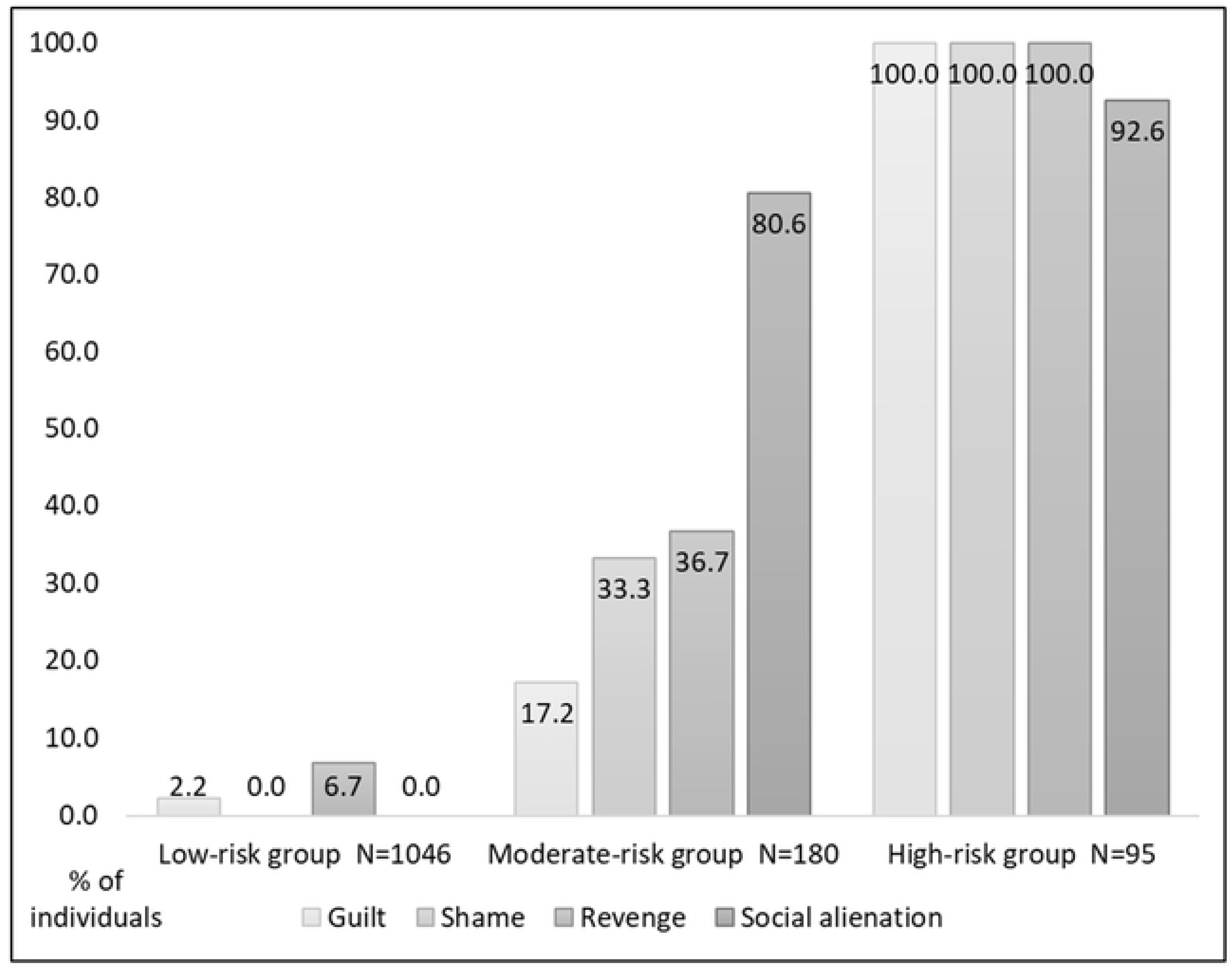
Percentage of individuals reporting the presence of guilt, shame, revenge and social alienation within each latent class of social-affective responses

**Table 5.**
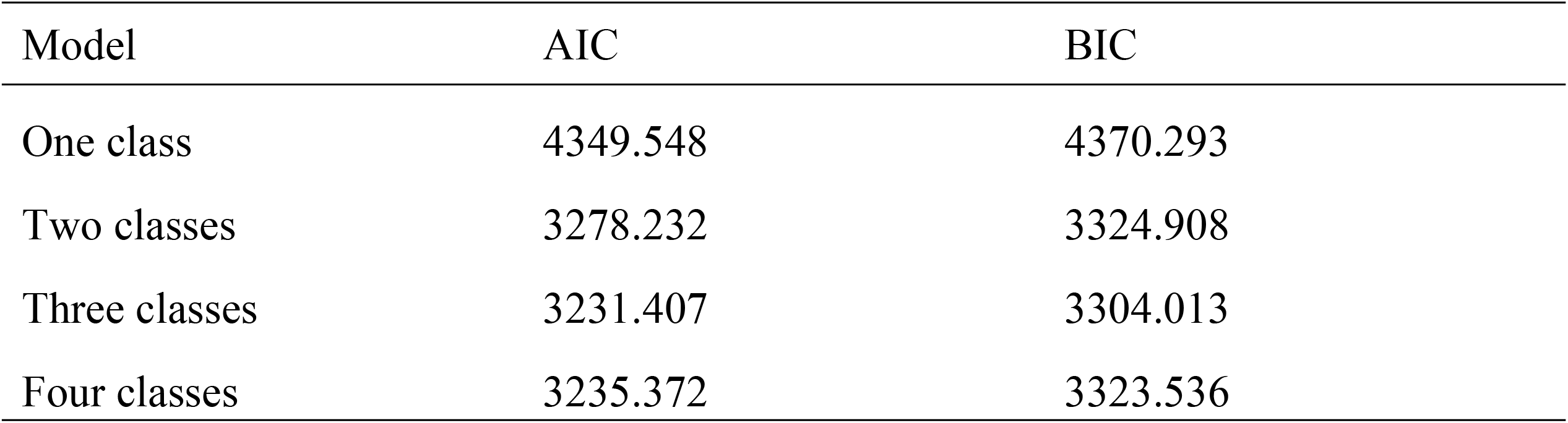
Results of Latent Class Analysis.

| Model | AIC | BIC |
| --- | --- | --- |
| One class | 4349.548 | 4370.293 |
| Two classes | 3278.232 | 3324.908 |
| Three classes | 3231.407 | 3304.013 |
| Four classes | 3235.372 | 3323.536 |

### Associations of class membership with mental disorders and with dimensional symptom measures (anxiety and depression)

Percentages of DD, PTSD, and AUD within the three latent classes are shown in Table 6. Descriptively, the highest percentage of PTSD (13.9%) and DD (11.1%) was in the moderate-risk group for social-affective responses, followed by the high-risk group (PTSD: 6.3%, DD: 6.3%) and the low-risk group for social-affective responses (PTSD: 2.2%, DD: 2.6%). In line with this, when compared to the low-risk group, the moderate-risk and the high-risk group for social-affective responses had a higher risk for PTSD (Moderate vs. Low: *OR* = 7.17, 95% CI = [3.97, 12.95], *p* < .001; High vs. Low: *OR* = 3.00, 95% CI = [1.19, 7.56], *p =*.020) and for DD (Moderate vs. Low: *OR* = 4.72, 95% CI = [2.58, 8.61], *p* < .001; High vs. Low: *OR* = 2.54, 95% CI = [1.02, 6.33], *p =*.044). There were no statistical differences between the moderate-risk and the high-risk group in the percentage of PTSD and DD (Table 6).

**Table 6.**
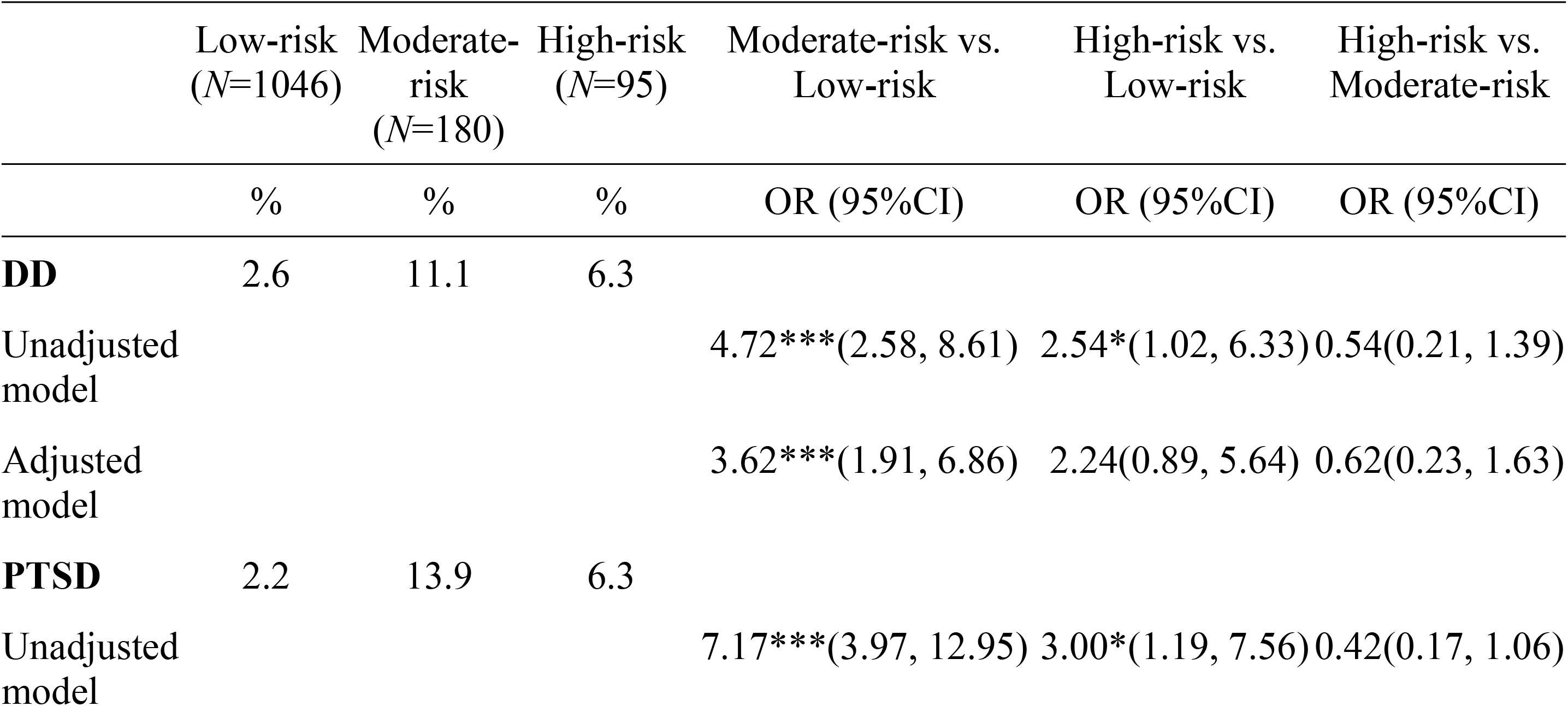
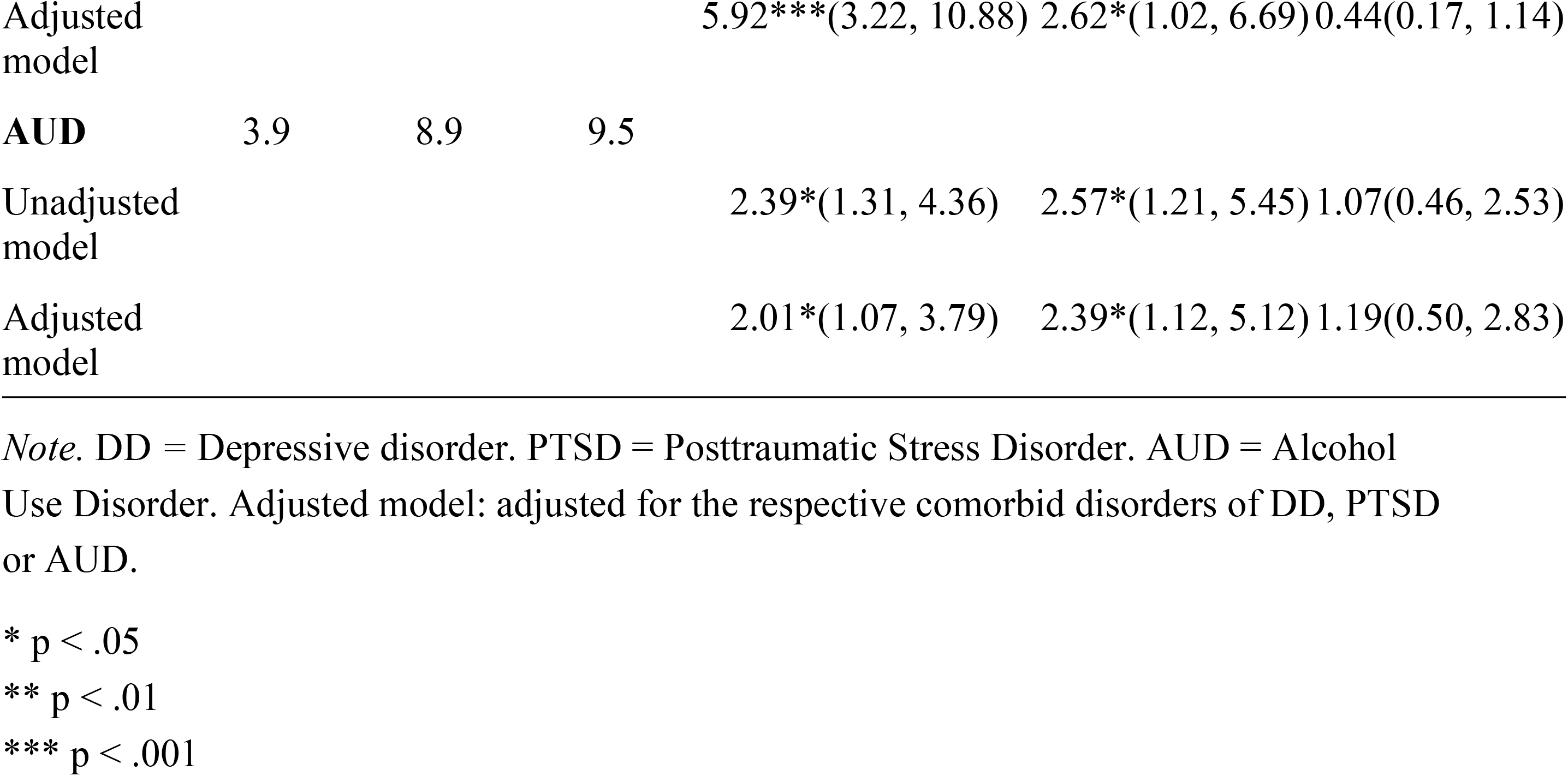
Percentage of DD, PTSD and AUD within each latent class of social-affective responses and associations between latent class membership and diagnoses.

|  | Low-risk<br>(*N*=1046) | Moderate-risk<br>(*N*=180) | High-risk<br>(*N*=95) | Moderate-risk vs.<br>Low-risk | High-risk vs.<br>Low-risk | High-risk vs.<br>Moderate-risk |
| --- | --- | --- | --- | --- | --- | --- |
|  | % | % | % | OR (95%CI) | OR (95%CI) | OR (95%CI) |
| **DD** | 2.6 | 11.1 | 6.3 |  |  |  |
| Unadjusted model |  |  |  | 4.72***(2.58, 8.61) | 2.54*(1.02, 6.33) | 0.54(0.21, 1.39) |
| Adjusted model |  |  |  | 3.62***(1.91, 6.86) | 2.24(0.89, 5.64) | 0.62(0.23, 1.63) |
| **PTSD** | 2.2 | 13.9 | 6.3 |  |  |  |
| Unadjusted model |  |  |  | 7.17***(3.97, 12.95) | 3.00*(1.19, 7.56) | 0.42(0.17, 1.06) |
| Adjusted model |  |  |  | 5.92*** (3.22, 10.88) | 2.62* (1.02, 6.69) | 0.44 (0.17, 1.14) |
| **AUD** | 3.9 | 8.9 | 9.5 |  |  |  |
| Unadjusted model |  |  |  | 2.39* (1.31, 4.36) | 2.57* (1.21, 5.45) | 1.07 (0.46, 2.53) |
| Adjusted model |  |  |  | 2.01* (1.07, 3.79) | 2.39* (1.12, 5.12) | 1.19 (0.50, 2.83) |

*Note.* DD = Depressive disorder. PTSD = Posttraumatic Stress Disorder. AUD = Alcohol Use Disorder. Adjusted model: adjusted for the respective comorbid disorders of DD, PTSD or AUD.
\* $p < .05$
\*\* $p < .01$
\*\*\* $p < .001$

With regard to the percentage of AUD, a slightly different pattern emerged: descriptively, the high-risk group for social-affective responses had the highest percentage of AUD (9.5%), followed by the moderate-risk group (8.9%) and the low-risk group (3.9%). In line with this, the high-risk group (*OR* = 2.57 ,95% CI = [1.21, 5.45], *p = .*014) and the moderate-risk group for social-affective responses (*OR* = 2.39, 95% CI = [1.31, 4.36], *p = .*004) had a higher risk for AUD than the low-risk group. The high-risk group and the moderate-risk group did not differ from each other with respect to the percentage of AUD (Table 6). Adjusting for comorbid disorders did not considerably change the described pattern of results (Table 6).

Dimensional measures of anxiety and depressive symptoms for each latent class of social-affective responses are presented in Table 7. Similar to what was found for DD and for PTSD, the moderate-risk group descriptively had the highest mean values for depressive symptoms (*M* = 3.9) and for anxiety symptoms (*M* = 4.8), followed by the high-risk group and the low-risk group (Table 7). In accordance with this, the moderate-risk group (β = 2.09, 95% CI = [1.67, 2.52], *p* < .001) and the high-risk group (β = 1.36, 95% CI = [0.80, 1.93], *p* < .001) had higher anxiety symptoms than the low-risk group for social-affective responses. Moreover, the high-risk group for social-affective responses had lower anxiety symptoms than the moderate-risk group (β = -0.73, 95% CI = [-1.40, -0.06], *p* = .032).

**Table 7.** Dimensional symptom measures of anxiety and depression in each latent class of social affective responses and associations between latent class membership and dimensional symptom measures.

|  | Low-risk<br>(N=1046) | Moderate-risk<br>(N=180) | High-risk<br>(N=95) | Moderate-risk vs.<br>Low-risk | High-risk vs.<br>Low-risk | High-risk vs.<br>Moderate-risk |
| --- | --- | --- | --- | --- | --- | --- |
| | *M (SD)* | *M (SD)* | *M (SD)* | $\beta$ (95%CI) | $\beta$ (95%CI) | $\beta$ (95% CI) |
| **Depressive symptoms** | 1.9<br>(2.4) | 3.9<br>(3.3) | 3.4<br>(3.4) |  |  |  |
| Unadjusted model |  |  |  | 1.97*** (1.55, 2.38) | 1.51*** (0.95, 2.06) | -0.46 (-1.11, 0.20) |
| Adjusted model |  |  |  | 0.68*** (0.34, 1.02) | 0.67** (0.23, 1.11) | -0.01 (-0.52, 0.50) |
| **Anxiety symptoms** | 2.7<br>(2.5) | 4.8<br>(3.3) | 4.1<br>(3.5) |  |  |  |
| Unadjusted model |  |  |  | 2.09*** (1.67, 2.52) | 1.36*** (0.80, 1.93) | -0.73* (-1.40, -0.06) |
| Adjusted model |  |  |  | 0.84*** (0.50, 1.19) | 0.40 (-0.04, 0.85) | -0.44 (-0.96, 0.08) |

*Note.* *M* = Mean value. *SD* = Standard Deviation. Adjusted model: adjusted for depressive symptoms respectively anxiety symptoms.
\* $p < .05$
\*\* $p < .01$
\*\* $p < .001$

The moderate-risk group (β = 1.97, 95% CI = [1.55, 2.38], *p* < .001) and the high-risk group (β = 1.51, 95% CI = [0.95, 2.06], *p* < .001) also had higher depressive symptoms than the low-risk group. The moderate-risk and the high-risk group did not differ with respect to the magnitude of depressive symptoms (Table 7).

When adjusted for anxiety respectively depressive symptoms, all associations were reduced (Table 7), and the high-risk group did no longer differ from the low-risk and the moderate-risk group with respect to anxiety symptoms.

## Discussion

The first aim of the present study was to examine individual associations of social affective responses to trauma exposure (revenge, social alienation, guilt, shame) with categorical and dimensional indicators of psychopathology. The second aim was to investigate potential patterns of social-affective responses to trauma exposure and their relation to psychopathology.

All social-affective responses were related to a higher risk for all examined mental disorders (PTSD, DD, AUD) as well as to higher levels of depressive and anxiety symptoms. Interestingly, for both DD and PTSD, as well as for depressive and for anxiety symptoms, the strongest associations were observed with social alienation. So far, social alienation in response to trauma exposure has received relatively little attention. A meta-analysis from 2020 found only nine studies that investigated associations between trauma-related alienation and PTSD symptoms, but suggested a large effect size (30). Among those nine studies, two studies compared trauma-related fear, anger, betrayal, shame, self-blame and alienation with respect to different psychological symptoms (20, 37). One study found that, when investigated together, only alienation predicted PTSD and depressive symptoms (20) and the other study demonstrated that trauma-related alienation was the only variable that predicted all forms of investigated trauma-related distress (PTSD, dissociation, and depression symptoms) across different samples (37).

In the present study, the strong association between social alienation and PTSD might partly be explained to the fact that social alienation overlaps with the DSM-IV PTSD criterion “feeling of detachment or estrangement from others” (38). However, it seems unlikely that the association was attributable to this overlap alone, as trauma-related social alienation also most strongly predicted DD, anxiety symptoms and depressive symptoms. Trauma-related social alienation could contribute to psychopathology as it could interfere with an individual’s sense of identity, foster insecure attachment styles and associated emotional distress (20, 37) and lead to a reduced capacity to benefit from potential social resources (16). However, a relationship in the opposite causal direction seems also conceivable, since individuals with a psychopathology of depression, anxiety or posttraumatic stress often suffer from diminished interest or pleasure, demonstrate avoidance behavior and experience stigma, which could all lead to social withdrawal and promote feelings and cognitions of social alienation. This could result in a vicious cycle in which social alienation fosters psychopathology and higher psychopathology in turn reinforces social alienation.

Besides social alienation, shame was the strongest predictor of PTSD, whereas guilt was the weakest predictor of PTSD. This is in line with previous studies demonstrating that shame is more strongly related to PTSD than guilt (18, 19). Shame might be more aversive than guilt, because it does not only refer to one’s perceived misbehavior in a specific situation (e.g. “I did something bad”), but to more global negative self-appraisals (e.g. “I am bad ”) as well as to the perception of being devalued in the eyes of others (19). In the present study, trauma-related guilt appeared to be of particular relevance for DD, which may be partly due to the fact that excessive or inappropriate guilt is a potential symptom of major depressive disorder. Revenge was the strongest predictor of AUD. Contrary to shame and guilt, revenge has received very little attention as a social-affective response to trauma exposure, although interpersonal aggression is common among trauma survivors (5). Our findings highlight the importance of identifying not only self-critical responses to trauma exposure (e.g., shame, guilt) but also hostile reactions towards others.

Besides investigating individual associations between social-affective responses and psychopathology, the second aim of this study was to examine patterns of social-affective responses and their relation to psychopathology. Three latent classes were identified that fitted the data best reflecting groups with low, moderate and high risk for negative social affective responses. The found classes seem to primarily reflect the overall proneness to experience negative social-affective responses. There appear to be few systematic patterns of social-affective responses with a high risk for one social-affective response and a low risk for other social-affective responses. Therefore, individuals who are more prone to self-critical social-affective responses (e.g. guilt, shame) also seem to be more prone to report hostile reactions (e.g. revenge) and to report social alienation. This is consistent, for example, with theories assuming that shame can result in externalization of blame and anger towards others as well as in social withdrawal (15). It is also in line with theories suggesting that feelings and cognitions of revenge often activate shame and guilt (39).

In the present study, one exception was that in the moderate-risk group, social alienation was reported with high likelihood, whereas the risk of reporting other social affective reactions was considerably smaller. After trauma exposure, the threshold to experience social alienation might therefore be relatively low. One might also speculate that reporting social alienation is less stigmatized than reporting revenge, guilt, or shame.

As could be expected, the low-risk group for social-affective responses had the lowest risk for PTSD, AUD and DD and the lowest levels of depressive and anxiety symptoms. A more surprising finding was that the high-risk group did not show higher levels of psychopathology than the moderate-risk group for social-affective responses. In contrary, the high-risk group even had lower anxiety symptoms than the moderate-risk group. A possible explanation could be that the moderate-risk and the high-risk group differed not only in terms of the likelihood with which individuals in these groups reported social-affective responses, but also in the way they coped with distressing feelings and thoughts. It is conceivable that some individuals in the moderate-risk group relied more heavily on avoidant coping strategies (e.g. rumination, experiential avoidance, thought suppression) to down-regulate the experience of negative social-affective responses. Such avoidant strategies, however, are related to higher levels of internalizing and distress-related psychopathology, such as symptoms of PTSD, depression and anxiety (40, 41). Another explanation could be that, in the present study, social alienation was particularly relevant for psychopathology, and individuals in the moderate-risk and in the high-risk group differed little in the likelihood with which they reported social alienation. Taken together, it appears necessary to consider not only the mere presence of social-affective responses but also their regulation and other potentially relevant moderating factors to understand the relationship between social affective responses and psychopathology.

This study has several limitations. (1) We examined a relatively healthy sample with an average low frequency of self-reported negative social-affective responses and low levels of psychopathology. This is a limitation in three regards. First, it reduces the variance in the variables under investigation, which could have led to an underestimation of group differences or associations. Second, it leads to limited generalizability to populations with higher levels of social-affective responses and symptomatology. Third, social-affective responses were operationalized as dichotomous variables due to their low variance, leading to a loss of information compared to a dimensional measure. (2) We examined a male, military sample, which limits the generalizability of the findings. There is also a chance of underreporting of mental health problems in this sample (27) (3) There were no validated instruments available to assess all of the examined trauma-related social-affective responses. Despite careful theoretical considerations, the validity of the used items remains unclear. (4) This was a cross-sectional study, so no definite conclusions can be made about the temporal sequence of the variables studied. Longitudinal studies are needed to investigate the relationship between trauma-related social-affective responses and subsequent psychopathology.

Despite these limitations, several important implications can be drawn from the findings of the present study. Our results indicate that trauma-related social alienation, shame, guilt, and revenge are likely phenomena in individuals who meet criteria for AUD, DD and PTSD as well as in individuals with higher levels of depressive and anxiety symptoms. This is important since previous research suggests that negative social-affective responses contribute to a higher severity and to the maintenance of psychopathology (10, 17). In addition, it has been demonstrated that trauma-related shame, guilt and alienation are associated with poorer outcomes in exposure based treatments (16, 42) and that within-person change in trauma-related shame and guilt predict changes in psychopathology during treatment (42). This underlines the importance of considering social-affective responses as possible treatment targets. More specifically, individuals experiencing negative social affective responses could particularly benefit from cognitive interventions that challenge dysfunctional trauma interpretations (16, 43). Additionally, emotion-focused interventions aimed at promoting (self-)compassion represent a promising approach for individuals experiencing self-critical responses such as shame and guilt after trauma exposure (43). As these interventions also aim to enforce social connectedness, they might also be valuable for individuals experiencing social alienation. Moreover, individuals who feel socially alienated after trauma exposure could benefit from interpersonal skills training.

Our findings further suggest that it is important for both researchers and clinicians to keep in mind that the presence of self-critical responses to trauma exposure (e.g. shame, guilt) is often accompanied by hostile responses (e.g. revenge) and social alienation. Similarly, individuals who present primarily with hostile responses towards others could at the same time have problems with reduced self-esteem (10) and may strongly experience shame and guilt. Therefore, it seems important to also assess those social-affective responses that may not be initially reported by patients, especially if these responses could be perceived as stigmatizing.

To further understand the potential causal pathways between trauma-related social affective responses and subsequent psychopathology, future studies should investigate the relationship between social-affective responses and mental disorders in prospective longitudinal studies, ideally with multiple assessments shortly after trauma exposure. Upcoming studies should also examine the extent to which findings of the present study can be replicated in different samples, including different demographic groups (high-risk groups vs. general population), different gender groups, and groups with higher levels of psychopathology and negative social-affective responses.

## Data Availability

The datasets generated and analyzed during the current study are not publicly available due to privacy restrictions (i.e., no informed consent about public availability of the raw data from the participants during data collection in 2010) and security concerns because of the specific population (German military personnel). The corresponding author can be contacted for data availability which will be checked with the funding agency.

## Acknowledgments

The study was logistically supported by the staff of the “Centre for Psychiatry and Posttraumatic Stress” in Berlin. Sabine Schönfeld, Clemens Kirschbaum and Hans-Ulrich Wittchen contributed to the planning of the former study program. Beyond the co-authors (Sebastian Trautmann and Judith Schäfer), Christin Thurau, Michaela Galle, Kathleen Mark and Anke Schumann were involved in the logistical handling.

## Supporting information captions

**Table S1 *Comparison of participants included versus excluded due to missing data***

**Table S2 *Distribution of items measuring guilt, revenge, shame and social alienation***

## References

1. Dohrenwend BP. Adversity, stress, and psychopathology. Oxford: Oxford University Press; 1998.

2. Breslau N, Davis GC, Peterson EL, Schultz L. Psychiatric Sequelae of Posttraumatic Stress Disorder in Women. Archives of General Psychiatry. 1997;54(1):81–7.

3. Bonanno GA. Uses and abuses of the resilience construct: Loss, trauma, and health related adversities. Social Science and Medicine. 2012;74(5):753–6.

4. Ozer EJ, Best SR, Lipsey TL, Weiss DS. Predictors of posttraumatic stress disorder and symptoms in adults: a meta-analysis. Psychological Bulletin. 2003;129(1):52.

5. Maercker A, Hecker T. Broadening perspectives on trauma and recovery: A socio interpersonal view of PTSD. European Journal of Psychotraumatology. 2016;7(1):29303.

6. Maercker A, Horn AB. A socio-interpersonal perspective on PTSD: The case for environments and interpersonal processes. Clinical Psychology & Psychotherapy. 2013;20(6):465–81.

7. Zeller M, Yuval K, Nitzan-Assayag Y, Bernstein A. Self-compassion in recovery following potentially traumatic stress: Longitudinal study of at-risk youth. Journal of abnormal child psychology. 2015;43:645–53.

8. DePrince AP, Zurbriggen EL, Chu AT, Smart L. Development of the trauma appraisal questionnaire. Journal of Aggression, Maltreatment & Trauma. 2010;19(3):275–99.

9. Kubany ES, Haynes SN, Abueg FR, Manke FP, Brennan JM, Stahura C. Development and validation of the trauma-related guilt inventory (TRGI). Psychological Assessment. 1996;8(4):428–44.

10. Gäbler I, Maercker A. Revenge phenomena and posttraumatic stress disorder in former East German political prisoners. The Journal of Nervous and Mental Disease. 2011;199(5):287–94.

11. Foa EB, Ehlers A, Clark DM, Tolin DF, Orsillo SM. The Posttraumatic Cognitions Inventory (PTCI): Development and Validation. Psychological Assessment. 1999;11(3):303–14.

12. Ehlers A, Clark DM. A cognitive model of posttraumatic stress disorder. Behaviour Research and Therapy. 2000;38(4):319–45.

13. McDevitt-Murphy ME, Zakarian RJ, Olin CC. Assessment of emotion and emotion related processes in PTSD. In: Tull MT, Kimbrel NA, editors. Emotion in Posttraumatic Stress Disorder: Academic Press; 2020. p. 3–41.

14. Tracy JL, Robins RW, Tangney JP. The self-conscious emotions: Theory and research: Guilford Press; 2007.

15. Lee DA, Scragg P, Turner S. The role of shame and guilt in traumatic events: A clinical model of shame-based and guilt-based PTSD. British Journal of Medical Psychology. 2001;74(4):451–46.

16. Ehlers A, Clark DM, Dunmore E, Jaycox L, Meadows E, Foa EB. Predicting response to exposure treatment in PTSD: The role of mental defeat and alienation. Journal of Traumatic Stress. 1998;11(3):457–71.

17. Cunningham KC. Shame and guilt in PTSD. In: Tull MT, Kimbrel NA, editors. Emotion in posttraumatic stress disorder: Elsevier; 2020. p. 145–71.

18. Bannister JA, Colvonen PJ, Angkaw AC, Norman SB. Differential relationships of guilt and shame on posttraumatic stress disorder among veterans. Psychological Trauma: Theory, research, practice, and policy. 2019;11(1):35.

19. Shi C, Ren Z, Zhao C, Zhang T, Chan SHW. Shame, guilt, and posttraumatic stress symptoms: A three-level meta-analysis. Journal of Anxiety Disorders. 2021;82.

20. Mitchell R, Hanna D, Brennan K, Curran D, McDermott B, Ryan M, et al. Alienation appraisals mediate the relationships between childhood trauma and multiple markers of posttraumatic stress. Journal of Child & Adolescent Trauma. 2020;13(1):11–9.

21. Kunst M. PTSD symptom clusters, feelings of revenge, and perceptions of perpetrator punishment severity in victims of interpersonal violence. International Journal of Law and Psychiatry. 2011;34(5):362–7.

22. Aakvaag HF, Thoresen S, Wentzel-Larsen T, Dyb G, Røysamb E, Olff M. Broken and guilty since it happened: A population study of trauma-related shame and guilt after violence and sexual abuse. Journal of Affective Disorders. 2016;204:16–23.

23. Kubany ES, Abueg FR, Owens JA, Brennan JM, Kaplan AS, Watson SB. Initial examination of a multidimensional model of trauma-related guilt: Applications to combat veterans and battered women. Journal of Psychopathology and Behavioral Assessment. 1995;14(4):353–76.

24. Tran HN, Lipinski AJ, Peter SC, Dodson TS, Majeed R, Savage UC, et al. The Association Between Posttraumatic Negative Self-Conscious Cognitions and Emotions and Maladaptive Behaviors: Does Time Since Trauma Exposure Matter? Journal of Traumatic Stress. 2019;32(2):249–59.

25. Wittchen HU, Schönfeld S, Thurau C, Trautmann S, Galle M, Mark K, et al. Prevalence, incidence and determinants of PTSD and other mental disorders: design and methods of the PID-PTSD+ 3 study. International Journal of Methods in Psychiatric Research. 2012;21(2):98–116.

26. Schönfeld S. Autobiographical memory changes in posttraumatic stress disorder (PTSD) (Doctoral dissertation): Philipps-Universität Marburg; 2006.

27. Johnson HP, Agius M. A Post-Traumatic Stress Disorder review: the prevalence of underreporting and the role of stigma in the Military. Psychiatria Danubina. 2018;30(suppl. 7):508–10.

28. Kim S, Thibodeau R, Jorgensen RS. Shame, guilt, and depressive symptoms: a meta analytic review. Psychological Bulletin. 2011;137(1):68.

29. Orth U, Montada L, Maercker A. Feelings of revenge, retaliation motive, and posttraumatic stress reactions in crime victims. Journal of Interpersonal Violence. 2006;21(2):229–43.

30. McIlveen R, Curran D, Mitchell R, DePrince A, O’Donnell K, Hanna D. A meta-analytic review of the association between alienation appraisals and posttraumatic stress disorder symptoms in trauma-exposed adults. Journal of Traumatic Stress. 2020;33(5):720–30.

31. Wittchen HU, Pfister H. DIA-X-Interviews: Manual für Screening-Verfahren und Interview; Interviewheft Längsschnittuntersuchung (DIA-X-Lifetime); Ergänzungsheft (DIA-X-Lifetime); Interviewheft Querschnittuntersuchung (DIA-X-Monate); Ergänzungsheft (DIA-X-Monate); PC-Programm zur Durchführung des Interviews (Längs und Querschnittuntersuchung); Auswertungsprogramm. Frankfurt: Swets and Zeitlinger; 1997.

32. Wittchen HU, Lachner G, Wunderlich U, Pfister H. Test-retest reliability of the computerized DSM-IV version of the Munich-Composite International Diagnostic Interview (M-CIDI). Social Psychiatry and Psychiatric Epidemiology. 1998;33(11):568–78.

33. Reed V, Gander F, Pfister H, Steiger A, Sonntag H, Trenkwalder C, et al. To what degree does the Composite International Diagnostic Interview (CIDI) correctly identify DSM-IV disorders? Testing validity issues in a clinical sample. International Journal of Methods in Psychiatric Research. 1998;7(3):142–55.

34. Conway CC, Forbes MK, South SC. A Hierarchical Taxonomy of Psychopathology (HiTOP) primer for mental health researchers. Clinical Psychological Science. 2022;10(2):236–58.

35. Herrmann C, Buss U, Snaith RP. Hospital Anxiety and Depression Scale-German Version (HADS-D). Bern: Hans Huber; 1995.

36. StataCorp. Stata Statistical Software: Release 15. College Station: TX: StataCorp LLC 2017.

37. DePrince AP, Chu AT, Pineda AS. Links between specific posttrauma appraisals and three forms of trauma-related distress. Psychological Trauma: Theory, Research, Practice, and Policy. 2011;3(4):430–41.

38. American Psychiatric Association. Diagnostic and Statistical Manual of Mental Disorders: DSM-IV-TR: 4th Edition Text Revision. Washington, DC: American Psychiatric Association; 2000.

39. Horowitz MJ. Understanding and ameliorating revenge fantasies in psychotherapy. American Journal of Psychiatry. 2007;164(1):24–7.

40. Miethe S, Wigger J, Wartemann A, Trautmann S. Posttraumatic stress symptoms and its association with rumination, thought suppression and experiential avoidance: A systematic review and meta-analysis. Journal of Psychopathology and Behavioral Assessment. 2023.

41. Aldao A, Nolen-Hoeksema S, Schweizer S. Emotion-regulation strategies across psychopathology: A meta-analytic review. Clinical Psychology Review. 2010;30(2):217–37.

42. Øktedalen T, Hoffart A, Langkaas TF. Trauma-related shame and guilt as time varying predictors of posttraumatic stress disorder symptoms during imagery exposure and imagery rescripting—A randomized controlled trial. Psychotherapy Research. 2015;25(5):518–32.

43. Kümmerle S, Hammerstein S, Helmig EM, Müller-Engelmann M. K-METTA: Kognitive Techniken und Metta-Meditationen zur Reduktion traumabezogener Schuld-und Schamgefühle–Darstellung der Intervention anhand von zwei Fallbeispielen. Verhaltenstherapie. 2022;32(4):196–210.

